# Evaluation of Large Language Models in Thailand’s National Medical Licensing Examination

**DOI:** 10.1101/2024.12.20.24319441

**Authors:** Prut Saowaprut, Romen Samuel Rodis Wabina, Junwei Yang, Lertboon Siriwat

## Abstract

Advanced general-purpose Large Language Models (LLMs), including OpenAI’s Chat Generative Pre-trained Transformer (ChatGPT), Google’s Gemini and Anthropic’s Claude, have demonstrated capabilities in answering clinical questions, including those with image inputs. The Thai National Medical Licensing Examination (ThaiNLE) lacks publicly accessible specialist-confirmed study materials. This study aims to evaluate whether LLMs can accurately answer Step 1 of the ThaiNLE, a test similar to Step 1 of the United States Medical Licensing Examination (USMLE). We utilized a mock examination dataset comprising 300 multiple-choice questions, 10.2% of which included images. LLMs capable of processing both image and text data were used, namely GPT-4, Claude 3 Opus and Gemini 1.0 Pro. Five runs of each model were conducted through their application programming interface (API), with the performance assessed based on mean accuracy. Our findings indicate that all tested models surpassed the passing score, with the top performers achieving scores more than two standard deviations above the national average. Notably, the highest-scoring model achieved an accuracy of 88.9%. The models demonstrated robust performance across all topics, with consistent accuracy in both text-only and image-enhanced questions. However, while the LLMs showed strong proficiency in handling visual information, their performance on text-only questions was slightly superior. This study underscores the potential of LLMs in medical education, particularly in accurately interpreting and responding to a diverse array of exam questions.

## 1. Introduction

Large Language Models (LLMs) have demonstrated remarkable capabilities in medical question-answering (QA) tasks, showcasing their ability to comprehend medical terminology, understand clinical contexts, and generate responses to a wide array of medical queries [1, 2]. Specifically, these models have shown impressive performance on multiple-choice research benchmarks [3, 4, 5] by utilizing extensive datasets [6, 7, 8] and sophisticated algorithms [9, 5] due to their ability to learn from a diverse range of medical scenarios. Their ability to process and interpret both text and image data positions them at the forefront of artificial intelligence (AI)-driven clinical decision support, highlighting their potential to transform healthcare delivery and improve patient outcomes.

LLMs have shown remarkable potential in answering medical examinations, including those from the United States Medical Licensing Examination (USMLE) [10, 11, 12]. These exams require a deep understanding of medical concepts, clinical reasoning, and the ability to apply knowledge in practical scenarios. For instance, Kung et al. [10] found that LLMs achieved an accuracy rate of 87% on USMLE Step 1 questions while Gilson et al. [11] reported that GPT-3 reached 85% accuracy on a set of USMLE-style questions. Furthermore, LLMs have shown proficiency in various languages and settings since they have been effective in Germany [13], Peru [14], Belgium [15], Japan [16, 17, 18, 19], Taiwan [20], Chile [21], South Korea [22], and many other countries [23]. Additionally, LLMs have exhibited potential in Thailand, particularly in other examinations such as pathology [24].

Medical examinations often include image inputs [25, 26], which require the use of LLMs that can process both text and images. These models, known as multi-modal image-text-to-text models, are essential for such tasks [27, 28]. Since these models are quite modern, only a minority of previous studies were able to utilize them in their research [10, 29]. At the time of writing, the leading closed-source multi-modal LLMs are developed by prominent companies, including OpenAI’s ChatGPT, Google’s Gemini, and Anthropic’s Claude. These advanced LLMs integrate visual and textual information to understand and generate responses accurately [30, 31, 32]. For example, in a multiple-choice QA task, these models can analyse the accompanying images to extract relevant details, combine these visual data with the textual information, and then provide accurate answers based on a comprehensive understanding of both inputs.

The Thai National Licensing Medical Examination (ThaiNLE) lacks publicly accessible, specialist-confirmed study materials. Students often rely on recollections of exam questions remembered by their seniors from previous years, sharing and collaboratively answering these questions to the best of their ability [33]. This informal and fragmented approach to exam preparation is particularly challenging for students in less popular or more rural medical schools, who may have access to fewer of these resources, creating an unfair advantage for their peers in more urban and well-known institutions [34]. This disparity underscores the need for reliable, standardized study resources to ensure comprehensive and equitable access to high-quality educational materials for all medical students. In this study, we aim to assess the feasibility of using LLMs to answer questions from Step 1 of the ThaiNLE. By exploring the capabilities of these advanced models, we seek to determine their effectiveness in accurately interpreting and responding to the exam’s diverse range of questions.

## 2. Methods

### 2.1 Dataset

We utilized a mock examination dataset consisting of 300 multiple-choice questions in English [35]. The dataset includes general principles (G) and organ system (S) questions, organized into 20 groups, such as biochemistry and molecular biology, biology of cells, and human development and genetics (see Table 1). This mock examination is widely used and has a difficulty level comparable to real examinations, albeit simpler than Step 1 of the USMLE. Additionally, 10.2% of the questions included images. A total of 19 board-certified physicians from various specialties, including internal medicine, pediatrics, and pathology, verified the answers.

**Table 1:** Topics covered in different groups.

| Group | Topic |
| --- | --- |
| G1 | Biochemistry and molecular biology |
| G2 | Biology of cells |
| G3 | Human development and genetics |
| G4 | Normal immune responses |
| G5 | Pathogenesis, pathophysiology, basic pathological process & lab investigation |
| G6 | Gender, ethnic, and behavioral considerations affecting disease treatment and prevention, including psycho social, cultural, occupational, and environmental |
| G7 | Multisystem processes |
| G8 | General pharmacology |
| G9 | Quantitative methods |
| S2 | Hematopoietic and Lymphoreticular Systems |
| S3 | Central and Peripheral Nervous Systems |
| S4 | Skin and Related Connective Tissue |
| S5 | Musculoskeletal System and Connective Tissue |
| S6 | Respiratory System |
| S7 | Cardiovascular System |
| S8 | Gastrointestinal System |
| S9 | Renal/Urinary System |
| S10 | Reproductive System and Perinatal Period |
| S11 | Endocrine System |

### 2.2 Passing Scores

The original examination consists of 300 questions, covering topics that adhere to the *Medical Competency Assessment Criteria for National License 2012* [36]. We have renamed the topic identifiers to enhance clarity and avoid confusion. The original naming convention used “B1.x” to denote General Principle topics and “B2-B11” for Systems topics. To make this distinction clearer, we have renamed these categories to “G” for General Principle topics and “S” for Systems topics.

We used the national passing scores from the *Center for Medical Competency Assessment* as benchmark performance against LLMs. Table 2 presents the passing scores and national averages for the main (summer) examinations from 2019 to 2024. The mean passing score for the main exam rounds between 2019 and 2024 was 52.3%, while the national average was 56.1%. The passing scores show some fluctuations over the six-year period, with the highest percentage recorded in 2021 at 53% and the lowest in 2024 at 51%. The national average scores also exhibit variability, with a notable increase in 2024, reaching 63.05%, which is significantly higher than in previous years. The standard deviation ranges from 12.60 in 2020 to 16.08 in 2024, indicating varying degrees of score dispersion across the years.

**Table 2:** Passing scores and national averages for main (summer) exams.

| Year | Passing Score (%) | National Average (%) | Standard Deviation |
| --- | --- | --- | --- |
| 2019 | 52.33 | 55.40 | 13.30 |
| 2020 | 52.67 | 53.80 | 12.60 |
| 2021 | 53.00 | 56.10 | 14.10 |
| 2022 | 52.84 | 54.20 | 15.10 |
| 2023 | 51.67 | 53.79 | 14.38 |
| 2024 | 51.00 | 63.05 | 16.08 |

### 2.3 Models and Inference

In this study, we utilized LLMs capable of processing both image and text data, including GPT-4, Claude 3 (C3)-Opus, and Gemini (G) 1-Pro and 1.5-Pro. These models offer advanced capabilities in terms of token processing and multi-modal input handling, making them suitable for complex tasks involving both textual and visual information. Both GPT-4 and C3-Opus can handle inputs of up to 32,768 tokens, enabling the efficient processing of large volumes of text data. Gemini-1.0-Pro and Gemini-1.5-Pro also support extensive token limits, though their exact capacities were not specified. These capabilities allow these models to manage and analyze large datasets effectively, integrating and synthesizing information from both text and images seamlessly. All models were accessed using the application programming interface (API) *requests* platform. We set the temperature to zero to provide deterministic outputs and minimize randomness from the models. Temperature is one of the parameters in LLMs that control the randomness of the model’s predictions, ranging from zero (deterministic) to 1 (diverse outputs). By opting for a lower temperature value, the model’s proclivity towards generating inventive outputs is curtailed, thereby providing predictability and consistency. The models were selected at each time point and their respective release dates are shown in Supplementary Table A1.

Each model predicted the answers to the entire dataset over a total of five inference runs. Questions were inputted one at a time to avoid confusing the model and to simulate how a student might input questions individually. The answers from each run were compared to the ground truth and scored accordingly. The scores from each run were then averaged, and a 95% confidence interval was calculated.

A zero-shot prompt was used that serves as the foundational instruction outlining the specific question and task to be executed by the LLMs. The prompt outputs a single letter to articulate clear directives for the model to follow in QA task. The specific prompt used was: “Given the following medical multiple-choice question, select the best answer from the provided options. Your response should be in the form of a single letter (A, B, C, D, E). Do not provide any explanation or additional information—only the letter or digit corresponding to the correct answer. Answer the following question:”

### 2.4 Evaluation Metrics

To evaluate the performance of the LLMs, we employed overall accuracy (micro-average) and balanced accuracy (macro-average), along with their 95% confidence intervals. Using both metrics provides a comprehensive evaluation of the LLMs’ performance. While micro-average gives an overall performance view, macro-average ensures that the performance on each class is equally considered, ensuring that no class is neglected.

Micro-average is determined by the percentage of correct answers, as shown in Equation 1. It emphasizes the overall performance by giving equal weight to each instance.

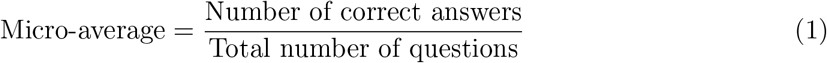

While micro-average is heavily influenced by the most frequent classes, we also utilized macro-average (Equation 2) since it gives equal weight to each subtopic, regardless of the number of instances in each subtopic, ensuring that the performance on all subtopics is considered equally.

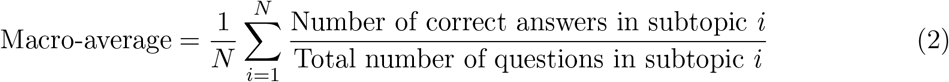

## 3. Results

Table 3 shows the overall and group accuracy of GPT, Claude, and Gemini models. The overall accuracy of the LLMs shows a clear hierarchical improvement led by GPT-4o that achieved the superior performance across all models at 88.9% (88.7, 89.1). It is followed by GPT-4 at 83.3% with a wider confidence interval between 79.6% and 87.0%, suggesting variability within its results. Meanwhile, Claude-3.5-Sonnet obtained 80.1% (79.6, 80.6), which is comparatively higher than its predecessor (Claude-3-Opus) which only obtained 77.8% (77.4, 78.2). The Gemini models achieved the poorest performance where Gemini-1.5-Pro only obtained 72.8% (72.2, 73.4) while Gemini-1.0-Pro acquired 61.4% (61.3, 61.5). In terms of macro-average, LLMs generated consistent results that also reflect similar trends and improvements across all models. The high balanced accuracy of GPT-4o at 89.1% (88.8, 89.4) indicates that it performs equally well across diverse topics, confirming its robustness and reliability in handling various data distributions.

**Table 3:** Overall Results.

| Model | Micro-average (%) | Macro-average (%) |
| --- | --- | --- |
| Gemini-1.0-Pro | 61.4 (61.3 - 61.5) | 63.1 (62.9 - 63.3) |
| Gemini-1.5-Pro | 72.8 (72.2 - 73.4) | 75.5 (75.0 - 76.0) |
| Claude-3-Opus | 77.8 (77.4 - 78.2) | 77.1 (76.9 - 77.3) |
| Claude-3.5-Sonnet | 80.1 (79.6 - 80.6) | 81.7 (81.5 - 81.9) |
| GPT-4 | 83.3 (79.6 - 87.0) | 82.8 (78.7 - 86.9) |
| GPT-4o | 88.9 (88.7 - 89.1) | 89.1 (88.8 - 89.4) |

**Table 4:** Micro-Accuracy Across Each Topic.

| Group | G1.0-Pro | G1.5-Pro | C3-Opus | C3.5-S | GPT-4 | GPT-4o |
| --- | --- | --- | --- | --- | --- | --- |
| G1 | 60.0 | 100.0 | 80.0 | 90.0 | 85.0 | 90.0 |
| G2 | 90.0 | 100.0 | 100.0 | 90.0 | 100.0 | 100.0 |
| G3 | 60.0 | 100.0 | 80.0 | 80.0 | 80.0 | 80.0 |
| G4 | 66.7 | 76.7 | 85.0 | 85.0 | 83.3 | 91.7 |
| G5 | 64.0 | 76.8 | 80.8 | 91.2 | 88.8 | 90.4 |
| G6 | 80.0 | 80.0 | 88.0 | 78.0 | 83.0 | 86.0 |
| G7 | 50.0 | 100.0 | 50.0 | 100.0 | 75.0 | 100.0 |
| G8 | 66.7 | 58.3 | 80.0 | 68.3 | 82.5 | 91.7 |
| G9 | 40.0 | 70.0 | 78.0 | 80.0 | 74.0 | 80.0 |
| S1 | 83.3 | 50.0 | 66.7 | 96.7 | 80.0 | 83.3 |
| S2 | 56.2 | 62.5 | 67.5 | 82.5 | 79.4 | 87.5 |
| S3 | 59.1 | 68.2 | 67.3 | 76.4 | 82.7 | 86.4 |
| S4 | 81.7 | 83.3 | 85.0 | 91.7 | 99.2 | 100.0 |
| S5 | 58.3 | 55.0 | 61.7 | 58.3 | 67.5 | 83.3 |
| S6 | 63.6 | 71.8 | 83.6 | 81.8 | 80.9 | 90.0 |
| S7 | 45.5 | 68.2 | 70.9 | 64.5 | 80.0 | 82.7 |
| S8 | 63.6 | 68.2 | 74.5 | 81.8 | 82.3 | 86.4 |
| S9 | 46.7 | 62.0 | 75.3 | 78.7 | 83.0 | 90.0 |
| S10 | 59.1 | 67.3 | 80.9 | 76.4 | 79.1 | 86.4 |
| S11 | 68.2 | 90.9 | 86.4 | 81.8 | 90.5 | 95.5 |

### 3.1 Results By Topic

Figure 1 presents the results for questions categorized under either General Principles or Systems. All models demonstrate higher accuracy on General Principle questions compared to Systems questions. The variability in scores for General Principles is relatively low, with most models achieving similar results. Specifically, the Gemini-1.0-Pro shows an accuracy of 64.1%, while the Gemini-1.5-Pro significantly improves to 84.6%. The Claude-3-Opus and Claude-3.5-Sonnet models further enhance accuracy, reaching 80.2% and 84.7%, respectively. The GPT-4 model achieves 83.5%, and GPT-4o leads with an impressive accuracy of 90.0%. In contrast, the Systems category exhibits more variability across models. The Gemini-1.0-Pro starts at 62.3%, with the Gemini-1.5-Pro improving to 67.9%. Claude-3-Opus and Claude-3.5-Sonnet show further gains with accuracies of 74.5% and 79.1%, respectively. The GPT-4 model achieves 82.2%, while GPT-4o again outperforms all others with an accuracy of 88.3%. Overall, GPT-4o consistently delivers the highest accuracy across both General Principles and Systems.

**Figure 1:**
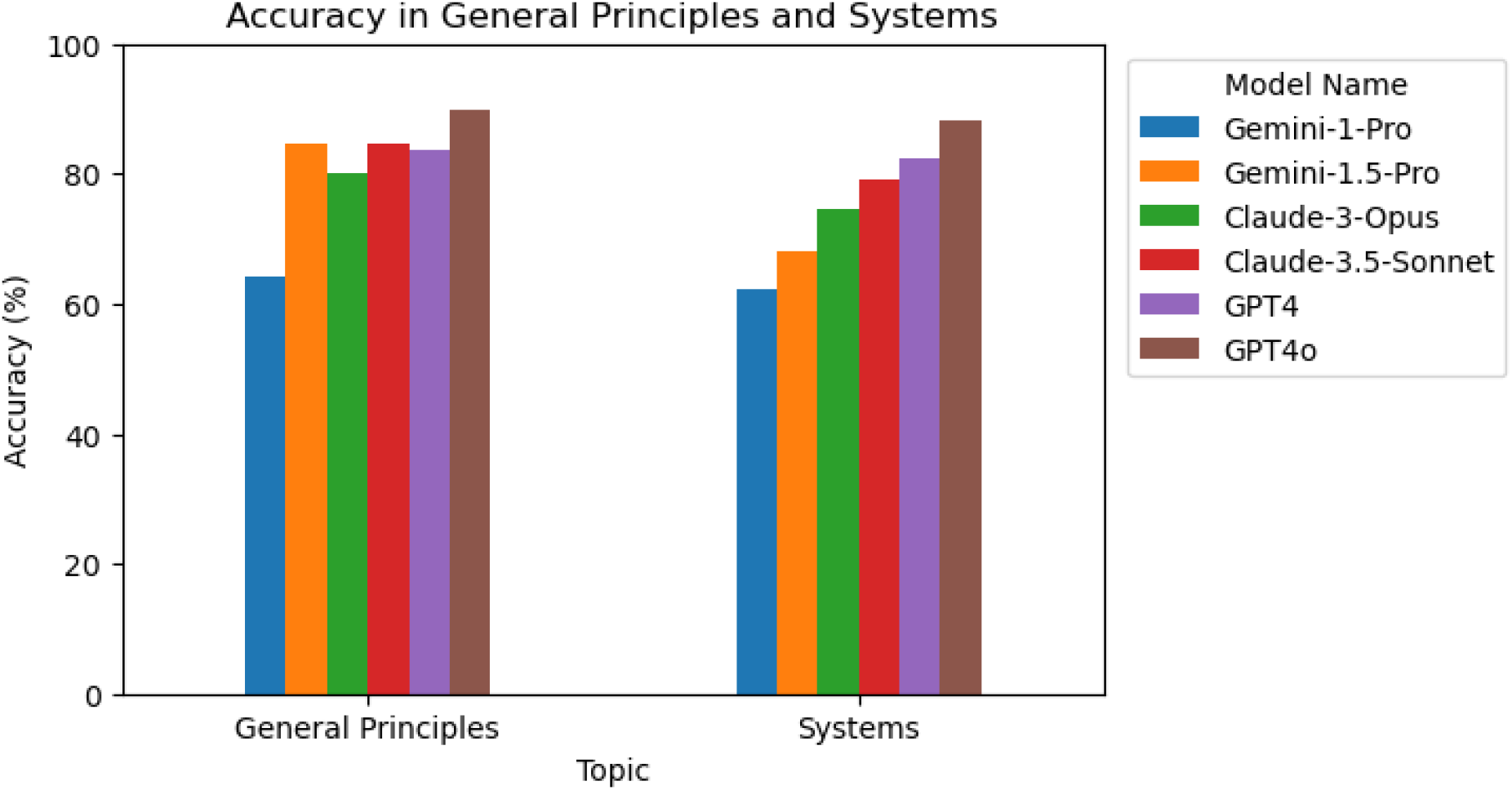
Macro-Accuracy in General Principles and Systems.

### 3.2 Comparing Questions With and Without Images

The comparative analysis of model accuracy for questions with and without images is detailed in this section, showcasing the performance variations across different models.

#### 3.2.1 Questions With Images

As illustrated in Table 5, the micro-accuracy for questions accompanied by images demonstrate a range of performances across the models. The Gemini-1.0-Pro achieves an accuracy of 70.3% (69.0, 71.6), while the Gemini-1.5-Pro shows a slightly lower accuracy of 68.4% (66.0, 70.8). Claude-3-Opus and Claude-3.5-Sonnet models further improve accuracy, achieving 72.9% (68.2, 77.6) and 74.8% (73.5, 76.1), respectively. The GPT-4 exhibits a higher accuracy of 78.4% (75.0, 81.8), with the GPT-4o reaching the highest accuracy of 82.6% (81.1, 84.1).

**Table 5:** Micro-Accuracy for Questions With Images.

| Model | With Images | Without Images |
| --- | --- | --- |
| Gemini-1-Pro | 70.3 (69.0 - 71.6) | 60.4 (60.4 - 60.4) |
| Gemini-1.5-Pro | 68.4 (66.0 - 70.8) | 73.3 (72.7 - 73.9) |
| Claude-3-Opus | 72.9 (68.2 - 77.6) | 78.4 (78.0 - 78.8) |
| Claude-3.5-Sonnet | 74.8 (73.5 - 76.1) | 80.7 (80.1 - 81.3) |
| GPT-4 | 78.4 (75.0 - 81.8) | 83.8 (80.0 - 87.6) |
| GPT-4o | 82.6 (81.1 - 84.1) | 89.6 (89.4 - 89.8) |

Most models performed better on questions without images compared to those with images. The comparison is shown in Figure 2. Interestingly, the Gemini-1.0-Pro model performed better on image questions, with an accuracy increase of 9.9 percentage points. The GPT-4 and GPT-4o models consistently outperformed others in both categories, with GPT-4o achieving the highest accuracy overall. The variation in performance between image and non-image questions suggests that while some models handle visual inputs effectively, others may not be as robust.

**Figure 2:**
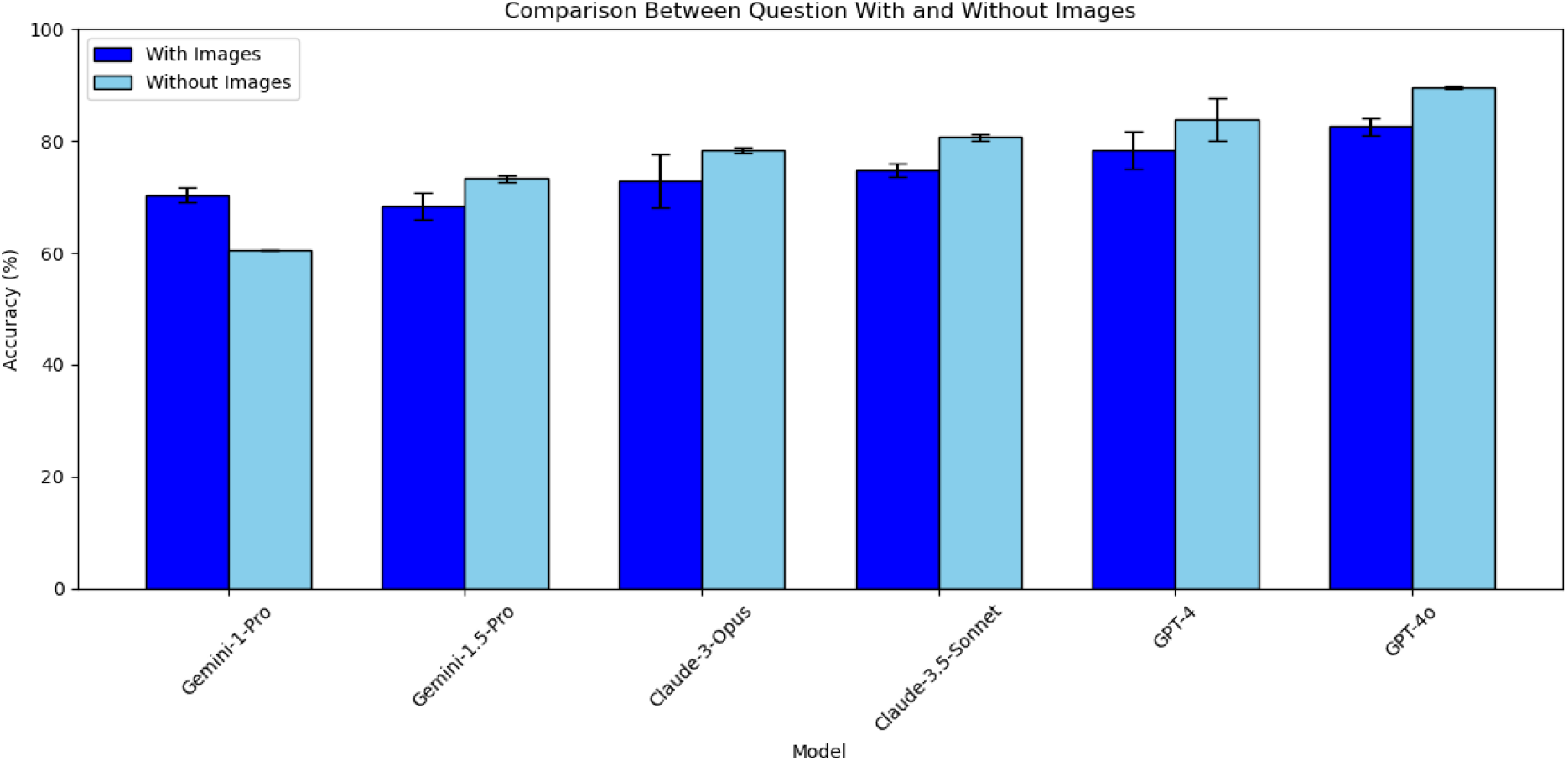
Comparison Between Question With and Without Images.

## 4. Discussion

In this work, we evaluated GPT, Gemini, and Claude in the ThaiNLE using a mock examination dataset comprising 300 multiple-choice questions. The findings highlight the significant potential of using LLMs for the ThaiNLE, effectively processing both textual and visual information. All models exceeded the passing score for all years. The LLMs demonstrated comparable performance across all topics, showcasing their versatility and robustness. They also showed proficiency with questions containing images, though there remains room for improvement to fully match the performance on text-only questions.

The trend observed with GPT models indicates a continuous improvement in accuracy, especially GPT-4o, which consistently outperformed all Claude and Gemini Pro models across all levels. These results corroborate prior studies demonstrating GPT-4o’s superior performance over Claude and Gemini in all sub-topics. For instance, [37] showed that GPT-4o achieved higher accuracy in Gastroenterology using the 2022 American College of Gastroenterology self-assessment examination compared to Gemini-Pro. Other studies also confirmed the superiority of GPT-4o in QA-tasks from specialized domains, such as ophthalmology, surgery, hypertension, and glaucoma management [38, 39, 40]. Nonetheless, all LLMs surpassed the passing score suggesting that future models, especially those trained or fine-tuned with specialized medical data, could offer even higher levels of precision. The integration of such models in medical examinations could revolutionize the way these tests are conducted and evaluated.

GPT, Claude, and Gemini models exhibit varying performance levels across different topics, excelling in some areas more than others. GPT-4 notably performed poorly on human development and genetics (G3) and quantitative methods (G9), while Claude struggled with the cardiovascular systems. In contrast, Gemini achieved the lowest performance in the musculoskeletal system and connective tissue (S5). According to the GPT-4 Technical Report, GPT-4 performs well on many benchmarks, but its performance on quantitative methods is notably weaker compared to other topics [41]. Meanwhile, Claude’s challenges with cardiovascular systems align with general observations that LLMs often have subject-specific strengths and weaknesses based on their training data and fine-tuning processes [42]. Specific studies on Claude’s performance across different medical domains are less documented in the open literature, indicating a need for further targeted research. The discrepancies in performance among these models may be due to differences in training data sources and fine-tuning methods [43, 44], although this warrants further investigation in future studies.

The performance of LLMs on text-based questions is generally superior to their performance on questions that include images. This phenomenon is consistent across all models except for the Gemini-1.0-Pro, which achieved a notable 70.3% accuracy on questions with images, outperforming its text-only performance. Despite being multi-modal models, GPT and Claude exhibited a discrepancy in handling visual information effectively compared to text. This could be due to the models’ training processes, where text data often predominates, resulting in better proficiency in text-based tasks. This is evident in previous studies where they have concluded that text-based representations generally resulted in higher accuracy than image-based questions [45]. Another analysis explored the capabilities of vision-enabled LLMs in handling image-based emotion recognition tasks. It was found that while these models are proficient, their performance still lags behind their text-based capabilities, suggesting that the integration of visual information remains a complex challenge for LLMs [46]. Overall, the discrepancy in performance can be attributed to the predominance of text data in the training processes of these models [47]. Despite being multi-modal, many LLMs have been trained primarily on large text corpora, resulting in better proficiency with text-based tasks [45, 46]. However, models like Gemini-1-Pro demonstrate that with optimized training on multi-modal data, significant improvements in handling image-based questions are possible.

Despite the promising results demonstrated in this study, several limitations should be addressed. First, we were unable to obtain permission to make the original dataset publicly available, which poses a limitation on the reproducibility of this study. Additionally, the dataset contained a relatively small number of image-based questions, which may have constrained the comprehensive evaluation of the models’ multi-modal capabilities. Finally, this study focused exclusively on existing general-purpose LLMs, such as GPT, Claude, and Gemini, without exploring the potential advantages of fine-tuning these models on domain-specific medical data. Future research should investigate whether such fine-tuning could result in significant improvements in accuracy and domain-specific expertise.

In the future, there is promising potential for the development of open-source models that are freely available such as Qwen [48] and Llama [49]. As technology progresses, it is anticipated that these models will become more compact, enabling them to run efficiently on local devices such as standard laptops or smartphones [50]. This accessibility could democratize the availability of advanced AI tools, allowing students from various backgrounds to benefit from high-quality educational resources. Enhanced accessibility to these tools could significantly improve the equity of medical education, providing all students with equal opportunities to succeed regardless of their geographical or economic constraints. [51]

Furthermore, the use of LLMs in medical education could extend beyond examinations since these models could be employed to generate practice questions [52], provide detailed explanations for complex questions [53], and translate questions using accurate domain-specific terminology [54]. As AI continues to evolve, its role in education is likely to expand, offering new possibilities for enhancing the quality and accessibility of medical training by ensuring that students receive comprehensive, understandable, and contextually relevant study materials.

In conclusion, the implementation of LLMs for the ThaiNLE represents a significant step forward in leveraging AI to enhance educational outcomes. Continued advancements in AI technology, coupled with increased accessibility, hold the promise of transforming medical education and ensuring that all students have the tools they need to excel in their studies and future careers.

## Data Availability

Researchers seeking additional information may contact the corresponding author to discuss potential collaborations or restricted access under specific agreements.

## Conflicts of Interest

The authors declare no conflict of interest.

## Disclosure Statement

During the preparation of this work, the author(s) used ChatGPT in order to check grammar and wording of the manuscript. After using this tool/service, the author(s) reviewed and edited the content as needed and take(s) full responsibility for the content of the publication.

## Data Availability

The dataset analyzed during the current study is not publicly available due to restrictions imposed by the corresponding author and the authors responsible for the dataset. Sharing the data would require explicit permission from potentially all 19 specialists, a process that is not feasible within the scope of this study. As such, access to the data is restricted to safeguard the confidentiality and intellectual property rights of those involved. Researchers seeking additional information may contact the corresponding author to discuss potential collaborations or restricted access under specific agreements.

## Funding

This research received no external funding.

## Appendix

### A. Supplementary Information

**Table A1:** Release Dates of Various Models.

| Model | Release Date |
| --- | --- |
| Gemini 1.0 Pro Vision | 15/02/24 |
| Gemini 1.5 Pro | 24/05/24 |
| Claude 3 | 14/03/24 |
| Claude 3.5 Sonnet | 21/06/24 |
| GPT-4 | 14/03/23 |
| GPT-4o | 13/05/24 |

### B. Sample Questions

1. A 6-year-old girl presents with acute dysentery. Stool examination revealed the egg as shown in the picture. Which of the following are the most likely activities that lead to her infection?

**Figure B1:**
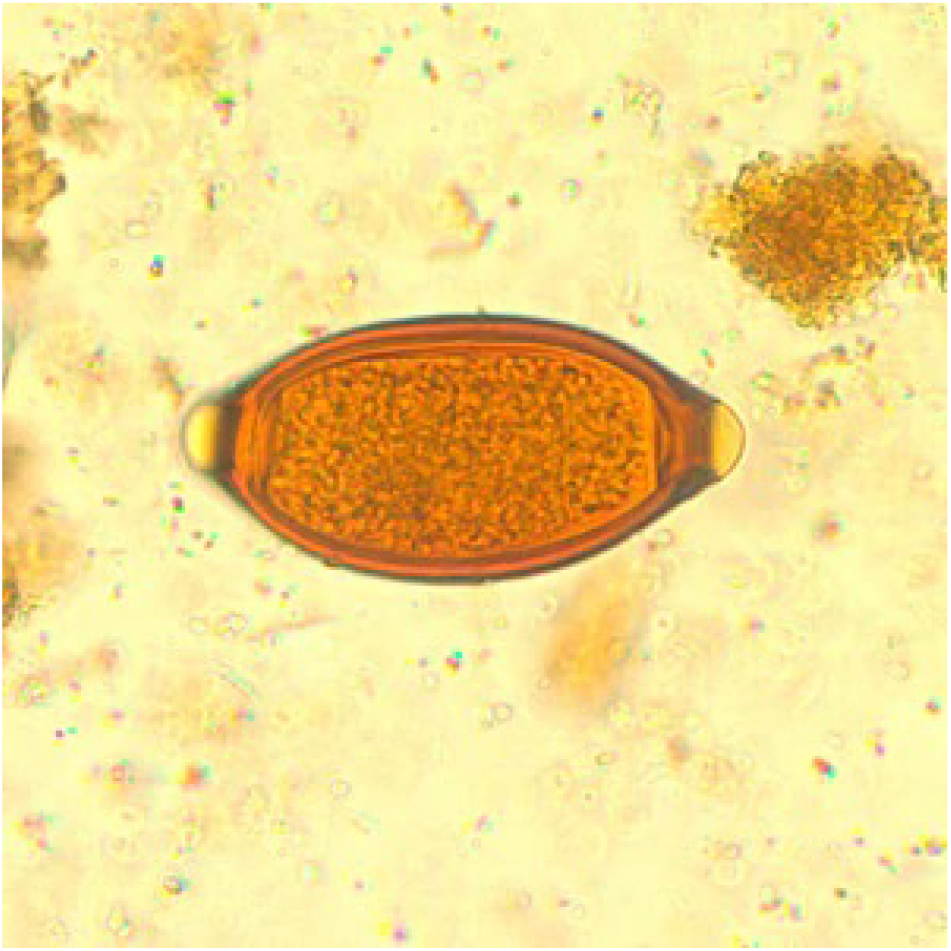
Trichuris trichiura Egg [55].

A. Eating raw fish
B. Barefoot walking
C. Eating uncooked pork
D. Eating fresh vegetables
E. Swimming in a dirty pond
2. A 37-year-old man was brought to the emergency department due to a motorcycle accident. Initial resuscitation was done, and his vital signs were stable. Physical examination revealed contusion at the right parietal area of the skull. However, his consciousness did not return until 3 hours later. His consciousness fully returned for about 8 hours, then he became comatose, arrested, and died a few minutes after his second loss of consciousness. According to this patient, what is the most likely finding in his autopsy?
  A. Accumulation of blood in the abdominal cavity
  B. Lens-shaped blood clot in the epidural space at the parietal area
  C. Concave-shaped blood in the subdural space
  D. Fracture of the skull base
  E. Brain edema
3. A 5-year-old girl presents with fatigue and jaundice for 2 days. Physical examination reveals pale conjunctivae, icteric sclerae, and splenomegaly. CBC shows Hb 8 g/dL, Hct 23%, MCV 65 fL, WBC 6,500 /µL (Neutrophil 80%, Lymphocyte 20%), platelet 300,000 /µL Her peripheral blood smear is as below: What is the most likely underlying defect of anemia?

**Figure B2:**
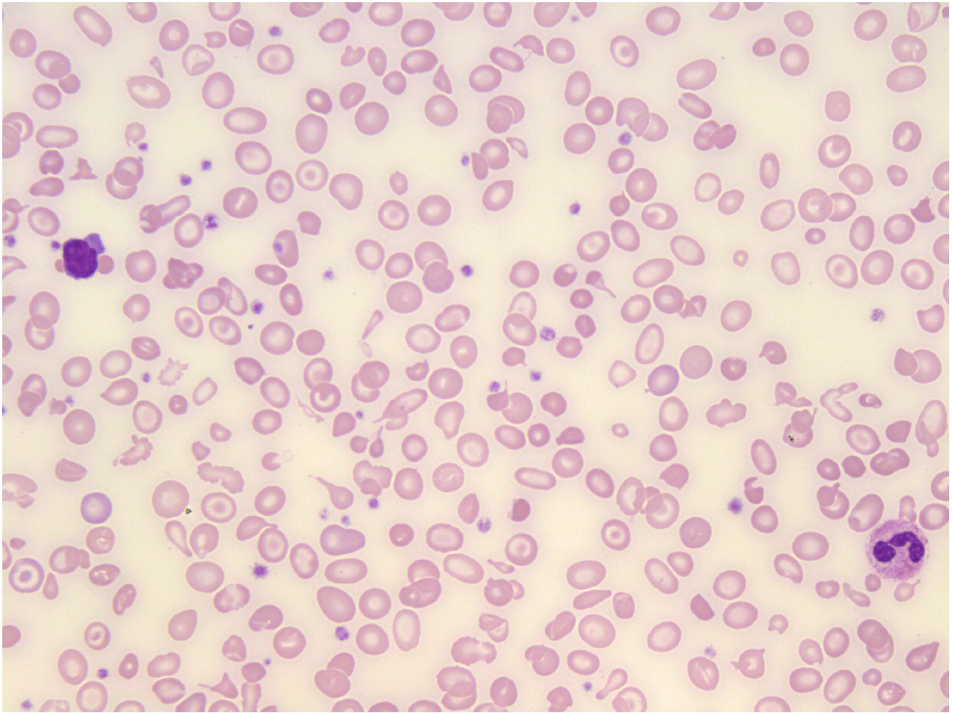
Peripheral Blood Smear of Thalassemia [56].

A. Abnormal globin chain production
B. Abnormal membrane cytoskeleton
C. Unstable heme production
D. Deficiency of red blood cell enzyme
E. Autoantibody to red blood cell

## Notes

### Competing Interest Statement

The authors have declared no competing interest.

### Funding Statement

The author(s) received no specific funding for this work.

### Author Declarations

Exempted–no humans involved

